# Trends in pregnancy-associated cervical cancer in Japan between 2012 and 2017: A multicenter survey

**DOI:** 10.1101/2022.02.06.22270505

**Authors:** Sayako Enomoto, Kosuke Yoshihara, Eiji Kondo, Akiko Iwata, Mamoru Tanaka, Tsutomu Tabata, Yoshiki Kudo, Eiji Kondoh, Masaki Mandai, Takashi Sugiyama, Aikou Okamoto, Tsuyoshi Saito, Takayuki Enomoto, Tomoaki Ikeda

## Abstract

Large-scale data on maternal and neonatal outcomes of pregnancy-associated cervical cancer in Japan are scarce, and treatment strategies have not been established. We conducted this multicenter retrospective observational study at 523 secondary/tertiary care hospitals in Japan to investigate the clinical features and trends in pregnancy-related cervical cancer treatments. We collected data for 290 patients with pregnancy-associated cervical cancer (during pregnancy, n=203; postpartum, n=87) diagnosed between January 1, 2012, and December 31, 2017. Of the 40 patients diagnosed at ≥22 gestational weeks, 34 (85.0%) were carefully followed until delivery without intervention. Of the 163 patients diagnosed at <22 gestational weeks, 111 and 52 patients continued and terminated their pregnancies, respectively. Although the termination rate increased with cervical cancer stage, 90 patients with stage IB1 disease had a variety of treatment options, including termination of pregnancy. When we divided the 59 stage IB1 patients who continued their pregnancy into four groups based on the primary treatment (strict follow-up, conization, trachelectomy, and neoadjuvant chemotherapy), there were no significant differences in progression-free or overall survival. The percentile of birth weight at delivery was smaller in the neoadjuvant chemotherapy group than in the strict follow-up group (*P* =.02). The full-term delivery rate was relatively higher in the trachelectomy group (35%) than in the other groups. In conclusion, treatment decisions, including pregnancy termination for pregnancy-associated cervical cancer, should be made after estimating the stage, with careful consideration of both maternal and fetal benefits. These findings will help in developing treatment guidelines for pregnancy-associated cervical cancer.

## 1. Introduction

Malignant disease in pregnancy occurs in approximately 1.0 per 1,000 deliveries worldwide.^1, 2^ In particular, the frequency of invasive cervical cancer in pregnancy is reported to be 1-12 per 10,000 pregnancies.^2-4^ Unfortunately, the incidence of pregnancy-associated cervical cancer is expected to increase in Japan because of the increasing proportion of births among women aged ≥35 years^5, 6^ and the increasing incidence of cervical cancer among younger women.^7^ In surveys of pregnant women with malignant disease conducted in 2008 and 2014, we identified cervical cancer as the most common type of cancer during pregnancy.^8, 9^ However, we could not investigate the trends in pregnancy-associated cervical cancer in Japan because both surveys were single-year studies and different facilities participated. Moreover, our previous studies focused on all types of malignancies during pregnancy and could not discuss clinical management for pregnancy-associated cervical cancer.

Previous studies have identified several aspects of pregnancy-associated cervical cancer; that is, pregnancy does not adversely affect the prognosis of cervical cancer^10, 11^, and chemotherapy during pregnancy results in small-for-gestational-age (SGA) newborns.^1, 12, 13^ Additionally, in 2009, a French Working Group proposed recommendations for the management of invasive cervical cancer in pregnant women. The International Network on Cancer, Infertility and Pregnancy (INCIP) also published guidelines for the management of gynecological cancers, including cervical cancer, in 2009 and has recently updated the guidelines.^14, 15^ Cervical cancer treatment guidelines in various countries also now include a chapter on pregnancy-associated cervical cancer.^4, 16, 17^ However, these guidelines for managing pregnancy-associated cervical cancer are not necessarily based on sufficient evidence. Indeed, there are no large-scale data on the long-term prognosis of pregnancy-associated cervical cancer. Therefore, this study aimed to clarify the incidence trends, treatment modalities, and maternal and neonatal outcomes in pregnancy-associated cervical cancer in Japan.

## 2. Materials and methods

### 2.1 Study design and patient population

This multicenter retrospective observational study was conducted at 523 secondary or tertiary care hospitals in Japan. In the primary survey, the number of patients with pregnancy-associated cervical cancer diagnosed between January 1, 2012, and December 31, 2017, was examined. In the secondary survey, clinicopathological information was retrospectively collected based on the medical records of the patients with pregnancy-associated cervical cancer. The exclusion criteria were as follows: missing data, pregnancy or postpartum not within the study period, twin pregnancy, and carcinoma *in situ*.

The study was conducted according to the tenets of the Declaration of Helsinki. Institutional review board approval was obtained from the Japan Society of Obstetrics and the Gynecology’s Clinical Research Committee (2018-7-2-76), Mie University School of Medicine (hosting institution, approval no. 3215), and all participating institutions. The study protocol was registered on the University Hospital Medical Information Network (UMIN 00003478). Informed consent was obtained in an opt-out form on the website.

### 2.2 Data collection and variable definitions

Clinical information collected in the secondary survey included age, history of pregnancy and delivery, diagnosis, primary stage, histology, gestational age at diagnosis, treatment modality, delivery method, additional treatment after delivery, and date of the last follow-up. Clinical staging was determined according to the 2008 International Federation of Gynecology and Obstetrics (FIGO) system. The percentile of birth weight was calculated based on the birth size standards by gestational age for Japanese neonates.^18^ SGA was defined as a birth weight of < the 10th percentile.

Pregnancy-associated cervical cancer was defined as cervical cancer histologically diagnosed during pregnancy, at delivery, or within 1 year postpartum. Histological findings included microinvasive carcinoma or worse, except for cervical intraepithelial neoplasia (CIN). Personal information of the participants was anonymized.

Japanese law prohibits abortions after 22 weeks of gestation. Therefore, we analyzed the clinicopathological features of cervical cancer during pregnancy separately in terms of diagnosis at <22 or ≥22 weeks of gestation. To identify the trends in treatment choice for cervical cancer during pregnancy by stage in Japan, we categorized the primary treatment modalities administered during pregnancy as follows: strict follow-up (delayed treatment after delivery), conization, trachelectomy, and neoadjuvant chemotherapy. When both conization and trachelectomy were performed, trachelectomy was considered the main treatment; for trachelectomy and adjuvant chemotherapy, trachelectomy was considered the main treatment; and for conization and neoadjuvant chemotherapy, neoadjuvant chemotherapy was considered the main treatment. The choices of equipment for conization, procedures for trachelectomy, and chemotherapy regimens depended on the criteria of each institution.

We focused on patients with stage IB1 disease who were diagnosed at a gestational age of <22 weeks because the treatment strategy for stage IB1 cervical cancer during pregnancy is controversial in the guidelines. We divided stage IB1 patients into the four groups based on the abovementioned primary treatment modalities during pregnancy, and we analyzed the impact of each treatment modality during pregnancy on both maternal and neonatal outcomes.

Overall survival was defined as the time from the date of diagnosis to the date of death or the last follow-up, and progression-free survival was defined as the time from the diagnosis date to disease progression.

### 2.3 Statistical analysis

The Cochran-Armitage trend test was used for the trend test. Continuous variables were compared between the four groups by one-way ANOVA and Tukey’s post hoc test. The Kaplan–Meier method and log-rank test were used to analyze progression-free and overall survival. All statistical analyses were performed using SPSS version 24.0 (IBM Corporation, Armonk, NY) or the R program (http://www.r-project.org). *P* <.05 was considered significant.

## 3. Results

### 3.1 Flow chart of this multicenter retrospective observational study

In the primary survey, 369 of 523 centers (72%) responded, of which 185 indicated that they recorded relevant cases of pregnancy-associated cervical cancer. In the secondary survey, 118 of 185 centers (64%) enrolled 307 patients (Figure 1). Of these 307 patients, 290 (203 patients diagnosed during pregnancy and 87 diagnosed postpartum) were included in the analysis according to the eligibility criteria.

**Figure 1.**
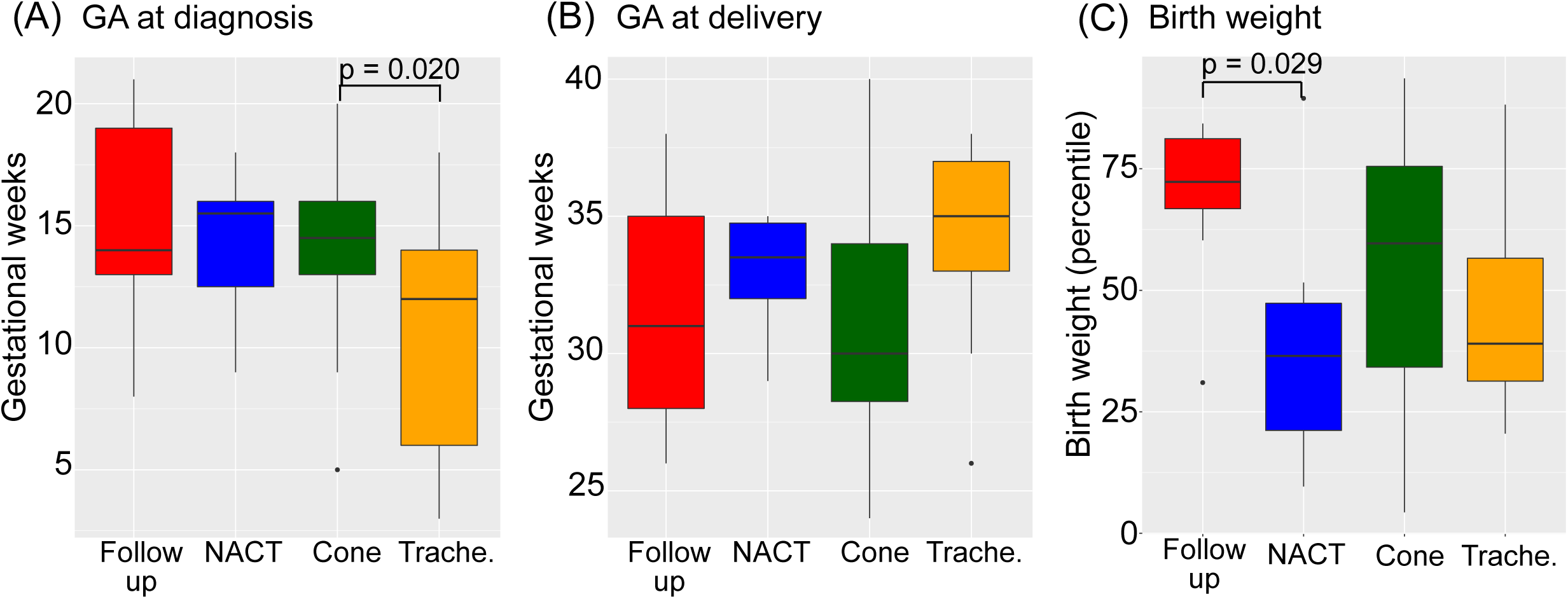
Patient inclusion flow chart.

### 3.2 Trends in pregnancy-associated cervical cancer

A total of 369,011 deliveries were recorded in the study from 2012 to 2017. When we defined the frequency of pregnancy-associated cervical cancer as the number of cases in each year divided by the total number of deliveries at all registered facilities, the frequency of cervical cancer during pregnancy (from 0.047% in 2012 to 0.068% in 2017, *P=*.037) showed a significantly increasing trend from 2012 to 2017 as shown in Figure S1.

### 3.3 Patient characteristics

Overall, 89% (260/290) of the patients had stage I disease (Table 1); among them, stage IB1 was the most common stage in both pregnancy (110/203 [54%]) and postpartum cases (37/87 [43%]). Among the patients diagnosed at ≥22 weeks of gestation, 15/40 (37.5%) had disease of the bulky type (i.e., tumor diameter ≥4 cm; stage IB2, IIA2), and the rate was significantly higher than that in the group diagnosed at <22 weeks of gestation (vs. 17/203 (8%), *P*<.001).

**Table 1.** Patient characteristics.

|  | Pregnancy cases<br>(n = 203) |  | Postpartum cases<br>(n = 87) |
| --- | --- | --- | --- |
|  | < 22 weeks (n = 163) | ≥22 weeks (n = 40) |  |
| Age at diagnosis (years) | 33.0 (31-36) | 32.0 (28-36) | 33.0 (29-37) |
| Nulliparous | 75 (46.0%) | 15 (37.5%) | 27 (31.0%) |
| Stage |  |  |  |
| IA1 | 40 (24.5%) | 3 (7.5%) | 19 (21.8%) |
| IA2 | 7 (4.3%) | 1 (2.5%) | 4 (4.6%) |
| IB1 | 90 (55.2%) | 20 (50.0%) | 37 (42.5%) |
| IB2-IVB | 26 (16.0%) | 16 (40.0%) | 27 (31.1%) |
| Histology |  |  |  |
| Squamous cell carcinoma | 114 (69.9%) | 20 (50.0%) | 68 (78.2%) |
| Adenocarcinoma | 39 (23.9%) | 13 (32.5%) | 13 (14.9%) |
| Others* | 10 (6.2%) | 7 (17.5%) | 6 (6.9%) |
| Gestational week at diagnosis | 14.0 (11-16) | 29.5 (27-34) | - |
| Number of months at diagnosis<br>after delivery | - | - | 2 (2-6) |

Data are presented as the median (IQR; range) or n (%). GA, gestational age; FIGO, International Federation of Gynecology and Obstetric.
\*Others include adenosquamous carcinoma, small-cell carcinoma, large-cell carcinoma, and neuroendocrine carcinoma.

The median gestational age at diagnosis was 14 (interquartile range [IQR]; range 11–16) weeks for the group diagnosed at <22 weeks of gestation and 29.5 (IQR; range 27–34) weeks for the group diagnosed at ≥22 weeks of gestation. In the group diagnosed at ≥22 weeks of gestation, 29/40 (72%) patients were diagnosed in the third trimester. The median time of diagnosis during the postpartum period was 2 (IQR; range 2–11) months after delivery.

There were no significant differences in maternal age at diagnosis, parity, or histology between the groups diagnosed at <22 and ≥22 weeks of gestation.

### 3.4 Pregnancy outcomes and treatment modality of the women diagnosed with cervical cancer at ≥22 weeks of gestation

Because artificial abortion is not an option in patients diagnosed at ≥22 weeks in Japan, all 40 patients continued their pregnancy and delivered. Among the patients diagnosed at ≥22 weeks of gestation, 85% (34/40) underwent strict follow-up, 5% (2/40) underwent neoadjuvant chemotherapy, and 10% (4/40) underwent surgical treatment via conization (n=3) or trachelectomy (n=1) (Table S1). Overall, 6/40 (15%) patients underwent treatment during pregnancy, and the median gestational age at diagnosis was 26.5 (IQR; range, 24–28) weeks. Of the 34/40 (85%) patients who did not undergo treatment during pregnancy, the median gestational age at diagnosis was 30 (IQR; range, 28–34) weeks. The median interval from diagnosis to delivery among the women who did not undergo treatment during pregnancy was 2 (IQR; range, 1–5) weeks (Figure S2), and 16/34 (47%) underwent concurrent radical hysterectomy and cesarean section.

### 3.5 Pregnancy outcomes and treatment modalities in the women diagnosed with cervical cancer at <22 weeks of gestation

The pregnancy outcomes by stage in the 163 patients diagnosed at <22 weeks of gestation are shown in Figure 2. The proportion of patients who opted for artificial abortions increased with stage progression, especially from stage 1B2 and above. In 47 patients who chose an artificial abortion, the median gestational ages at diagnosis and abortion were 12 (IQR; range, 9–15) and 16 (IQR; range, 12–19) weeks, respectively. Miscarriages occurred in 5/163 (3.1%) patients, 2 of which occurred after conization during pregnancy (surgeries at 15 and 19 weeks of gestation), 1 after trachelectomy (surgery at 17 weeks of gestation), and 2 among women who did not undergo treatment during pregnancy (Figure 1). The rate of live births tended to decline as the disease progressed.

**Figure 2.**
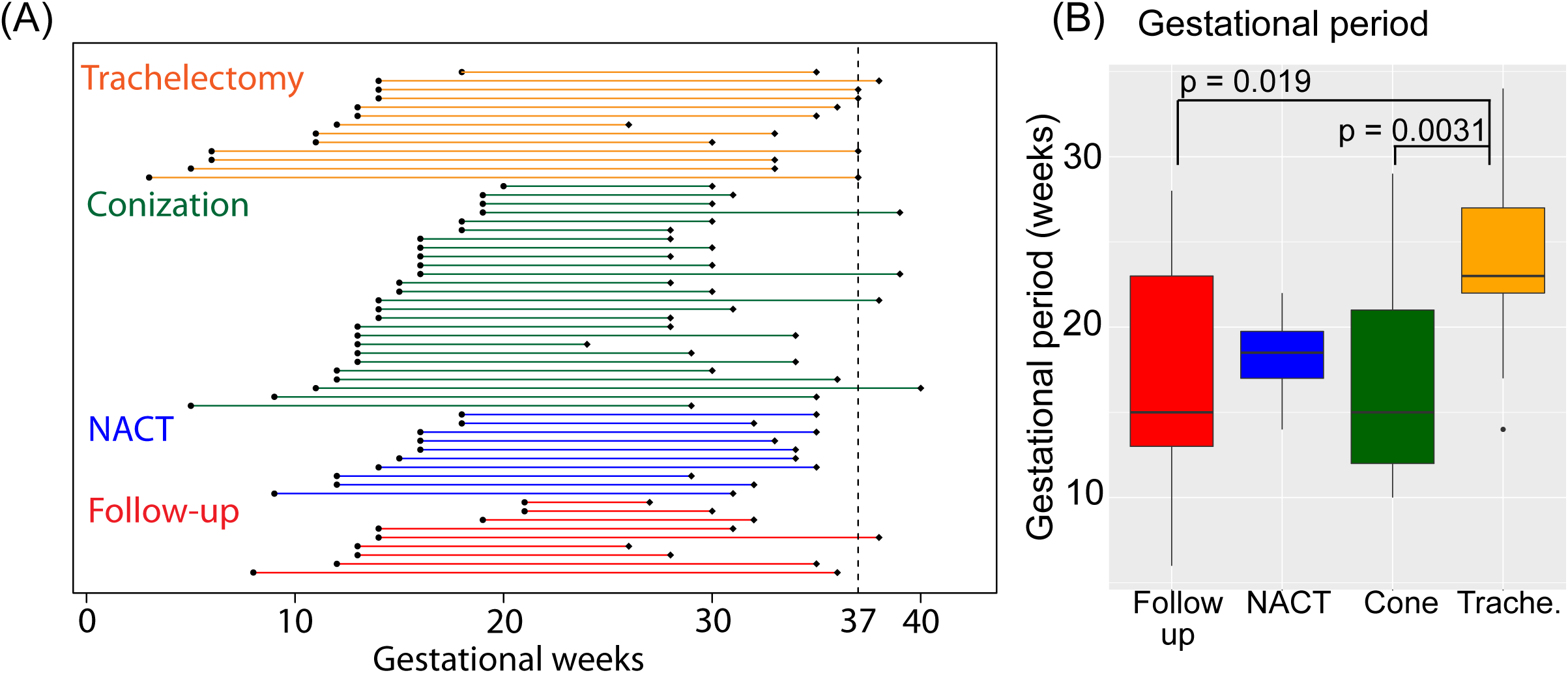
Outcomes during pregnancy by stage among women diagnosed at <22 weeks of gestation (n=163)

Next, treatments administered during pregnancy by stage among the patients who chose to continue their pregnancy before <22 weeks of gestation are shown in Table 2. Most women with cervical cancer (86.5%) were treated during their pregnancy. The main treatment modality was surgery, and 80/111 (72.1%) patients underwent conization or trachelectomy.

**Table 2.** Treatments administered during pregnancy by stage among patients diagnosed at a gestational age of <22 weeks who chose to continue their pregnancy (n = 111)

| Treatment during pregnancy | IA1<br>(n = 37) | IA2<br>(n = 5) | IB1<br>(n = 59) | IB2-IVB<br>(n = 10) |
| --- | --- | --- | --- | --- |
| Strict follow-up (n = 15) | 1 (2.7%) | 3 (60%) | 9 (15.3%) | 2 (20.0%)* |
| Conization (n = 64) | 35 (94.6%) | 2 (40%) | 26 (44.1%) | 1 (10.0%) |
| Trachelectomy (n = 16) | 1 (2.7%) | - | 14 (23.7%) | 1 (10.0%) |
| NACT (n = 16) | - | - | 10 (16.9%) | 6 (60.0%) |

Data are presented as n (%).
\*One person died of the disease, NACT; neoadjuvant chemotherapy.

In all, 35/37 (94.6%) patients with stage IA1 disease underwent conization, and there were no cases of maternal death during the observation period. The median gestational ages at diagnosis and delivery were 14 (IQR; 12-16) and 37 (IQR; 37-39) weeks, respectively. In the patients with stage IA1 disease treated with conization, 26/35 (74.3%) continued to full term, and 9/35 (25.7%) had preterm delivery (Figure S3). Among the 9 patients with preterm delivery, 3 underwent iatrogenic preterm delivery for the purpose of early treatment of cervical cancer. In contrast, 6/10 patients with stage IB2 or higher disease (60.0%) were treated with neoadjuvant chemotherapy, and 2 patients (20.0%) were followed up until ≥22 weeks of gestation. On the other hand, a wide variety of treatment modalities were selected by those with stage IB1 disease.

### 3.6 Treatment for stage IB1 disease diagnosed at <22 weeks of gestation

In 59 patients with stage IB1 cervical cancer, 26 (44.1%) underwent conization, 14 (23.7%) underwent trachelectomy, and 10 (16.9%) received neoadjuvant chemotherapy during pregnancy (Table 3). Nine patients were not treated with surgery or chemotherapy and were followed up until delivery. Table 3 shows the clinicopathological characteristics of each treatment modality during pregnancy. One patient who received adjuvant chemotherapy after trachelectomy was excluded from the subsequent analysis. The median gestational age at diagnosis tended to be earlier in the trachelectomy group than in the other groups, with a median of 12.5 weeks (IQR; range 7-14), and there was a significant difference between the two groups, when compared with conization (*P* =.020) (Figure 3A). Although trachelectomy was performed at a median of 16.0 weeks (IQR; range 15-17), neoadjuvant chemotherapy was started at a median of 19.5 weeks (IQR; range, 18-21) in consideration of the fetal organogenesis period. Although there was no significant difference in the gestational weeks at delivery among the four groups (Figure 3B), the percentage of full-term births tended to be higher in the trachelectomy group (35.8%) than in the strict follow-up group (11.1%), the conization group (15.4%), and the neoadjuvant chemotherapy group (0%). The cause of preterm birth was iatrogenic for the purpose of early treatment of cervical cancer in all cases. The percentage of patients who underwent surgery for the treatment of cervical cancer at the same time as cesarean section was 55.6% (5/9) in the strict follow-up group, 76.9% (20/26) in the conization group, 71.4% (10/14) in the trachelectomy group, and 100% (10/10) in the neoadjuvant chemotherapy group (Figure S4).

**Table 3.** Comparison of the clinicopathological data among the four groups of patients with stage IB1 cervical cancer diagnosed at <22 weeks of gestation.

|  | Strict follow-up<br>(n = 9) | Conization<br>(n = 26) | Trachelectomy<br>(n = 14) | NACT<br>(n = 10) |
| --- | --- | --- | --- | --- |
| Age at diagnosis (years) | 37.0 (31-39) | 34.0 (31-37) | 32.5 (29-34) | 36.0 (34-36) |
| Pathological tissue |  |  |  |  |
| Squamous cell carcinoma | 6 (66.7%) | 17 (65.4%) | 10 (71.4%) | 7 (70.0%) |
| Adenocarcinoma | 2 (22.2%) | 8 (30.8%) | 3 (21.4%) | 3 (30.0%) |
| Others | 1 (11.1%) | 1 (3.8%) | 1 (7.2%) | 0 (0%) |
| Gestational age<br>at diagnosis (weeks) | 14.0 (13-19) | 14.5 (13-16) | 12.5 (7-14) | 15.5 (13-16) |
| Gestational age<br>at first treatment (weeks) | - | 16.0 (15-19) | 16.0 (15-17) | 19.5 (18-21) |
| Gestational age<br>at delivery (weeks), | 31.0 (28-35) | 30.0 (28-34) | 35.0 (33-37) | 33.5 (32-35) |
| Preterm delivery | 8 (88.9%) | 22 (84.6%) | 9 (64.2%) | 10 (100%) |
| Full term delivery | 1 (11.1%) | 4 (15.4%) | 5 (35.8%) | 0 (0%) |
| Mode of delivery |  |  |  |  |
| Vaginal delivery | 1 (11.1%) | 2 (7.8%) | 0 (0%) | 0 (0%) |
| Cesarean section | 8 (88.9%) | 24 (92.2%) | 14 (100%) | 10 (100%) |
| Neonatal outcome |  |  |  |  |
| Birth weight percentile | 72.3 (67-81) | 60.0 (34-76) | 37.8 (31-53) | 36.5 (21-47) |
| Small for gestational age | 0 (0%) | 2 (7.8%) | 0 (0%) | 1 (10%) |
| NICU admission | 7 (77.8%) | 21 (80.8%) | 9 (64.2%) | 10 (100%) |

Data are presented as the median (IQR; range) or n (%)
NACT: neoadjuvant chemotherapy, NICU: neonatal intensive care unit

**Figure3.**
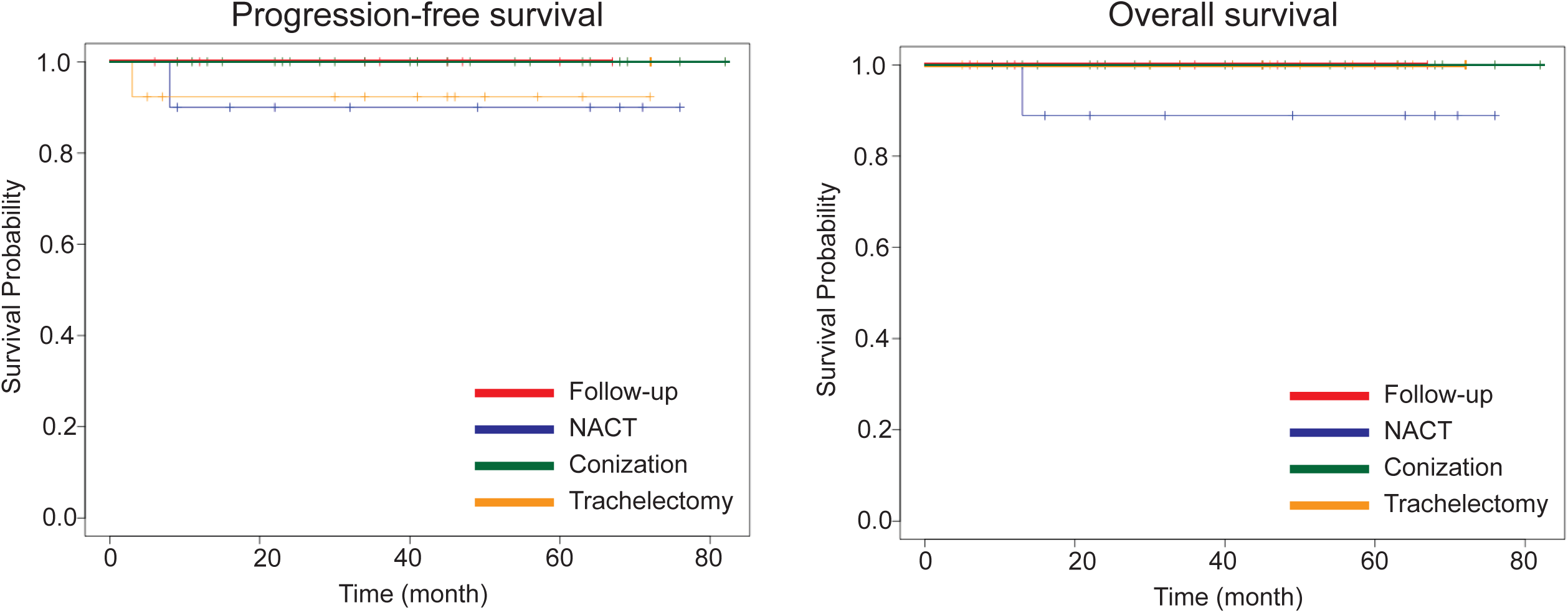
Comparison of gestational age at (A) diagnosis (B) delivery (C) birth weight percentile at delivery between the four groups.

For the infants, the birth weight percentile was 72.3 (IQR; range, 67-81) in the follow-up group. On the other hand, the birth weight percentile was 36.5 (IQR; range, 21-47) in the neoadjuvant chemotherapy group. It was significantly lower in the neoadjuvant chemotherapy group than in the follow-up group (*P* =.029) (Figure 3C). SGA was detected in 2/26 (7.8%) patients in the conization group and 1/10 (10.0%) patients in the neoadjuvant chemotherapy group.

Next, the duration of pregnancy for each treatment group was compared among the four groups (Figure 4). The duration for the trachelectomy group was significantly longer than those for the follow-up (*P* =.019) and conization groups (*P* =.0031).

**Figure4.**
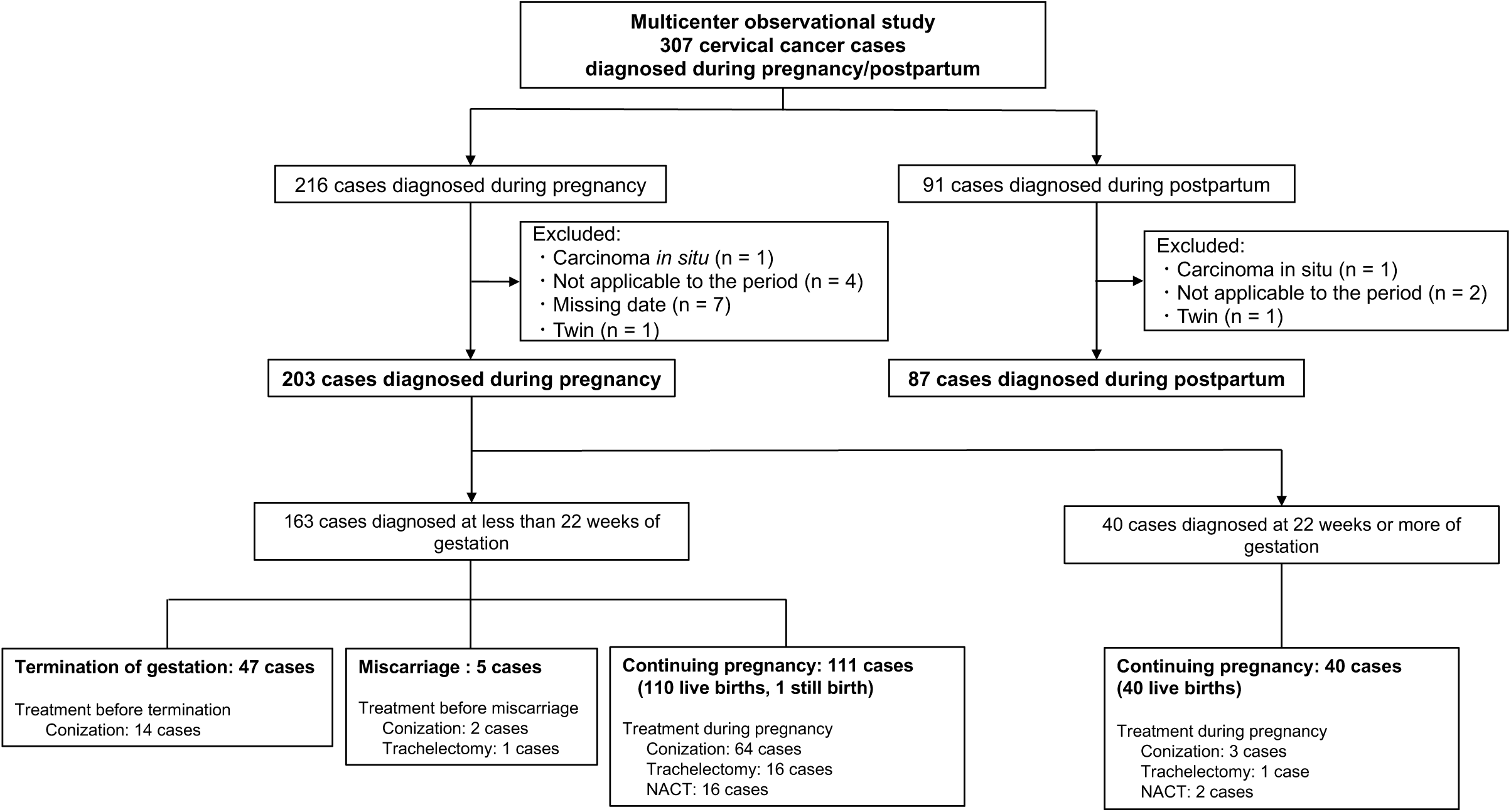
(A) Duration from diagnosis to delivery for all cases by the four groups. (B) Comparison of the duration of prolonged pregnancy between the four groups.

Finally, the oncologic outcome of each treatment modality was assessed by Kaplan–Meier survival analysis (Figure 5). No significant differences in progression-free or overall survival were observed among the four groups.

**Figure5.**
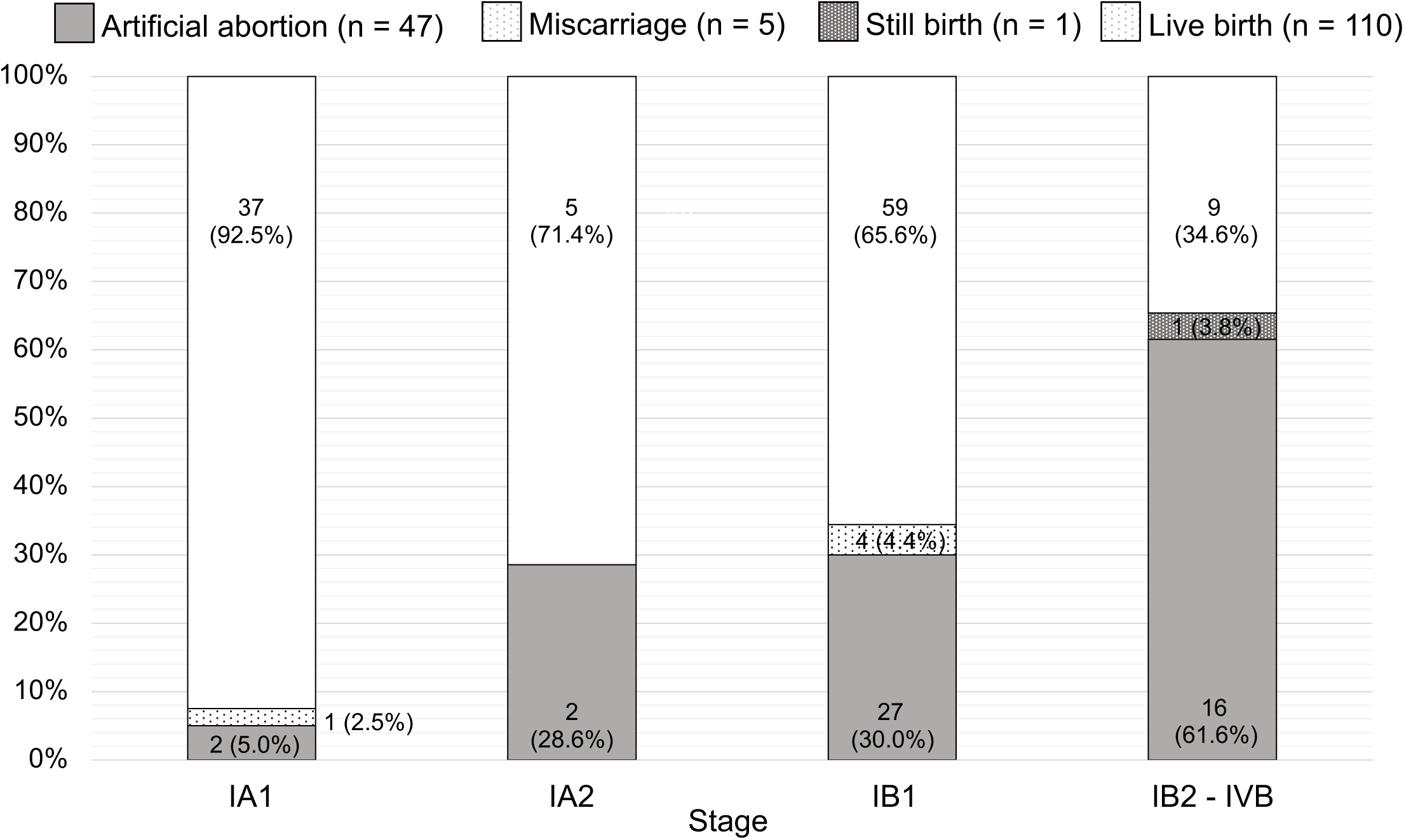
Comparison of oncologic outcomes between four groups.

## Discussion

Our multicenter retrospective observational study demonstrated that the frequency of cervical cancer during pregnancy increased between 2012 and 2017. The increasing trend of cervical cancer during pregnancy in Japan may be related to the decreased vaccination rate, which was due to the reported complications of the vaccine in Japan. Furthermore, the cervical cancer screening rate is lower in Japan than in other countries.^19, 20^ Thus, individuals should be encouraged to receive cervical cancer vaccination and cervical cancer screening.

In the Japanese guideline^4^ and the guideline of the International Consensus Meeting^15^ on the management of cervical cancer during pregnancy, conization is recommended as the treatment for stage IA disease without lymphovascular space invasion. In this study, 90% of the women with stage IA disease who continued pregnancy underwent conization, and no deaths occurred at the end of the study. These results support the recommendations of the guidelines. On the other hand, 17.6% of the patients with stage IA1 disease treated with conization during pregnancy had preterm births, except iatrogenic preterm births. A meta-analysis of retrospective studies of obstetric outcomes after conization in nonpregnant patients with CIN, including stage IA1 disease, reported that the preterm birth rate was 11.2%^21^. Although the number of stage IA1 patients was limited in this meta-analysis study, the risk for preterm birth in patients with stage IA disease treated with conization during pregnancy may be slightly higher than that in pregnant women treated with conization prior to becoming pregnant. Therefore, careful obstetric management is required after conization during pregnancy.

The greatest concern among physicians is the appropriate treatment modality for stage IB1 disease, which is the most common stage during pregnancy.^11, 22^ The French guideline^10^ and the guideline based on the third International Consensus Meeting^15^ have established a treatment plan for FIGO 2009 stage IB1 disease with a tumor size less than 2 cm. Both guidelines recommend pelvic laparoscopic lymphadenectomy for stage IB1 disease diagnosed before 22 weeks of gestation. If lymph node metastasis is positive, interruption of the pregnancy is recommended. If lymph node metastasis is negative, delayed treatment after delivery is recommended. The guideline based on the third International Consensus Meeting^15^ also recommends simple trachelectomy as another treatment option. The important point is that lymphadenectomy is not recommended after 22 weeks of gestation because a sufficient number of nodes cannot be retrieved after this gestational age.^15^

Although the French guideline^10^ was published in 2009, pelvic lymphadenectomy was not performed for stage IB1 disease diagnosed before 22 weeks of gestation in Japan between 2012 and 2017. The Japanese guideline^4^ states that therapeutic strategies should be discussed individually for patients with stage IB1 disease diagnosed before 22 weeks of gestation, and there are four treatment modalities (follow-up, neoadjuvant chemotherapy, conization, and trachelectomy) for stage IB1 disease. Although no significant difference in oncologic outcome was observed between the four treatment modalities, there were some differences in the gestational period and the birth weight percentile.

Specifically, the trachelectomy group tended to show an earlier diagnosis than the other groups. Abdominal trachelectomy is often performed at approximately 16 weeks to avoid miscarriage at an early gestational age and difficulties in surgical procedures at late gestational weeks.^23^ In other words, trachelectomy is a treatment modality for stage IB1 disease diagnosed at an early gestational age. The trachelectomy group also had a tendency to demonstrate a longer duration of pregnancy than the other groups. Indeed, the rate of full-term birth was higher in the trachelectomy group. This tendency might be influenced by the high radicality of trachelectomy compared to other treatment modalities. However, one miscarriage that occurred after trachelectomy was considered an iatrogenic abortion. Furthermore, the surgical techniques for trachelectomy during pregnancy are difficult to standardize and should thus be performed at high-volume centers. In addition, the neoadjuvant chemotherapy group showed lower birth weights, consistent with the findings of previous studies.^1, 12, 13^ However, no significant difference in the frequency of SGA among the four groups was observed, probably due to the small sample size of the neoadjuvant chemotherapy group. The frequency of SGA was significantly higher in the neoadjuvant chemotherapy group compared to the other groups (OR: 5.24, 95% CI: 1.24–22.1, *P=*0.024) when all cases were analyzed without limiting by stage (data not shown). A 20-year international cohort study of 1170 patients indicated that babies exposed to antenatal chemotherapy might be more likely to develop SGA and be admitted to the neonatal intensive care unit (NICU) than babies not exposed.^1^ Therefore, the involvement of hospitals with obstetric high-care units in the management of patients with invasive cervical cancer is recommended. In addition, the platinum agent frequently used for neoadjuvant chemotherapy is classified as group 2A by International Agency for Research Cancer (IARC), and is transferred to the fetus after maternal administration via placenta. Therefore, the long-term prognosis for the fetus (including carcinogenesis) should be carefully considered.

This study has some limitations because of its retrospective nature, utilization of data from case series (which comprised 60%), and utilization of data from selected hospitals in Japan. Due to the retrospective study, we were not able to update the stage of each case from the FIGO 2009 to the FIGO 2018.^24^ On the other hand, this is the first study to investigate pregnancy-associated cervical cancer and to reveal the trends in cervical cancer during pregnancy, including treatment strategies, in Japan. These results can be useful for developing management and treatment guidelines for cervical cancer during pregnancy. Moreover, the importance of early detection of cervical cancer through appropriate cervical cancer screening and the prevention of disease onset through active vaccination against cervical cancer was reiterated.

## Data Availability

All data created in the current work are included in the manuscript

## Abbreviations

FIGO: 2008 International Federation of Gynecology and Obstetrics
HR: hazard ratio
IQR: interquartile range
NICU: neonatal intensive care unit
OR: odds ratio
SGA: small for gestational age

## Acknowledgments

Our special thanks go to all the doctors of the following study facilities for their cooperation: Asahikawa-Kosei General Hospital, Sapporo City General Hospital, Sapporo Medical University Hospital, Hokkaido University Hospital, Iwate Medical University Hospital, Iwate Prefectural Central Hospital, Tohoku University Hospital, Fukushima Medical University Hospital, Kawaguchi Municipal Medical Center, University of Tsukuba Hospital, International University of Health and Welfare Hospital, Jichi Medical University Hospital, Gunma University Hospital, Saiseikai Kawaguchi General Hospital, Toho University Sakura Medical Center, Juntendo University Urayasu Hospital, Chiba University Hospital, Japanese Red Cross Medical Center, The Jikei University Hospital, The Cancer Institute Hospital Of Japan Foundation For Cancer Research, Tokyo Women’s Medical University Medical Center East, Teikyo University, Tokyo Women’s Medical University Hospital, The Jikei University Hospital, Nippon Medical School Hospital, Saiseikai Yokohamashi Tobu Hospital, Kameda Medical Center, National Hospital Organization Yokohama Medical Center, Yokohama City University Medical Center, Yokohama Rosai Hospital, Odawara Municipal Hospital, Chiba Cancer Center, Showa University Northern Yokohama Hospital, Kanagawa Children’s Medical Center, St. Marianna University School of Medicine, Tokai University Hospital, Fujisawa City Hospital, Niigata University Medical & Dental Hospital, Kanazawa University Hospital, Ina Central Hospital, Fukui University Hospital, Nagano Red Cross Hospital, Shinshu University Hospital, Gifu Prefectural Tajimi Hospital, Gifu Prefectural General Medical Center, Gifu University Hospital, Takayama Red Cross Hospital, Shizuoka City Shizuoka Hospital, Shizuoka Saiseikai General Hospital, Hamamatsu University Hospital, Aichi Cancer Center Hospital, Komaki City Hospital, Fujita Health University Hospital, Japanese Red Cross Nagoya Daiichi Hospital, Nagoya University Hospital, Nagoya City Universal Hospital, Shiga University of Medical Science Hospital, University Hospital Kyoto Prefectural University of Medicine, Kyoto University Hospital, National Hospital Organization Osaka National Hospital, Osaka City University Hospital, Osaka University Hospital, Saiseikai Suita Hospital, Higashiosaka City Medical Center, Kindai University Hospital, Kobe City Medical Center General Hospital, Kansai Rosai Hospital, Hyogo College of Medicine, Hyogo Cancer Center, Japanese Red Cross Wakayama Medical Center, Hashimoto Municipal Hospital, Tottori Prefectural Central Hospital, Tottori Municipal Hospital, Matsue City Hospital, Okayama Saiseikai General Hospital, Kurashiki General Hospital, Hiroshima Prefectural Hospital, Hiroshima City Hiroshima Citizens Hospital, JA Hiroshima General Hospital, JA Onomichi General Hospital, Hiroshima City Asa Hospital, National Hospital Organization Kure Medical Center, National Hospital Organization Iwakuni Clinical Center, Yamaguchi Prefectural Grand Medical Center, Yamaguchi University Hospital, Tokushima University Hospital, Takamatsu Red Cross Hospital, Ehime University Hospital, Kagawa Rousai Hospital, Tokuyama Central Hospital, Kochi Health Science Center, Kitakyushu Municipal Medical Center, Saiseikai Fukuoka General Hospital, Kyushu University Hospital, Kumamoto University Hospital, Kagoshima City Hospital, Nagasaki Harbor Medical Center, Nagasaki University Hospital, Miyazaki Prefectural Miyazaki Hospital, Miyazaki Prefectural Nobeoka Hospital, Saga University Hospital, Oita University Hospital, Miyazaki University Hospital, Kagoshima University Hospital, Okinawa Chubu Hospital, University of the Ryukyus Hospital, Iwate Prefectural Ninohe Hospital, Akita University Hospital, National Hospital Organization Fukuyama Medical Center, Fukuda Hospital, Ehime Prefectural Niihama Hospital, Yokohama City Minato Red Cross Hospital, Sasebo City General Hospital, Showa University Hospital, Osaka Police Hospital, Tsuboi Hospital, Tokyo Metropolitan Bokutoh Hospital, KKR Sapporo Medical center, Hokkaido Prefectural Esashi Hospital, Hirosaki University Hospital, Iwate Prefectural Chubu Hospital, Tohoku Medical and Pharmaceutical University Hospital, Omagari Kousei Medical Center, Ibaraki Seinan Medical Center Hospital, Tsukuba Medical Center Hospital, Sano Kosei Hospital, Chiba Rosai Hospital, Tokyo Metropolitan Health and Medical Corporation Toshima Hospital, Yokohama City University Medical Center, Nagaoka Red Cross Hospital, Ishikawa Prefectural Central Hospital, Sugita Genpaku Memorial Obama Municipal Hospital, Anjo Kosei Hospital, Yokkaichi Municipal Hospital, Japanese Red Cross Ise Hospital, Uji-Tokushukai Medical Hospital, Fukuchiyama City Hospital, Nara City Hospital, Japanese Red Cross Okayama Hospital, Fukuoka Tokushukai Hospital, Iizuka Hospital, Kumamoto Rosai Hospital, Almeida Memorial Hospital, Okinawa Red Cross Hospital, Asahikawa Medical University Hospital, Hakodate Municipal Hospital, Kushiro City General Hospital, Asahikawa City Hospital, Japanese Red Cross Asahikawa Hospital, Iwate Prefectural Ofunato Hospital, Iwate Prefectural Iwai Hospital, Iwate Prefectural Miyako Hospital, Sagamino Hospital, Chukyo Hospital, Hokkaido Hospital, Tochigi Medical Center, NTT Medical Center Tokyo, NTT Medical Center Sapporo, Aizenbashi Hospital, Kainan Hospital, Aidu Chuo Hospital, Ome Municipal General Hospital, Aomori Prefectural Central Hospital, Aomori City Hospital, Odate Municipal General Hospital, Akita Red Cross Hospital, Japanese Red Cross Ashikaga Hospital, Arao Municipal Hospital, Iida Municipal Hospital, Isesaki Municipal Hospital, Ichinomiya Municipal Hospital, Ibaraki Prefectural Central Hospital, Tsuchiya General Hospital, Iwamizawa Municipal General Hospital, Ehime Prefectural Central Hospital, Hokkaido P.W.F.A.C. Engaru-Kosei General Hospital, Oji General Hospital, Omihachiman Community Medical Center, Oita Prefectural Hospital, Oita Red Cross Hospital, Osaka International Cancer Institute, Osaka Red Cross Hospital, Osaka Women’s and Children’s Hospital, National Hospital Organization Osaka Minami Medical Center, Osaka Rosai Hospital, Osaki Citizen Hospital, Ohta General Hospital, Ohara General Hospital, Omuta City Hospital, Kagawa Prefectural Central Hospital, Kagawa University Hospital, Kagoshima Prefectural Oshima Hospital, Kasukabe Medical Center, Japanese Red Cross Tokyo Katsushika Perinatal Hospital, Kanagawa Cancer Center, Kanazawa Medical Center, Shinko Hospital, Kawasaki Municipal Hospital, Kansai Medical University Hospital, Kitasato University Hospital, Japanese Red Cross Kitami Hospital, Gifu Municipal Hospital, Kimitsu Chuo Hospital, National Hospital Organization Kyushu Cancer Center, Japanese Red Cross Kyoto Daiichi Hospital, Japanese Red Cross Kyoto Daini Hospital, Kyoto Chubu Medical Center, Kyoto Katsura Hospital, North Medical Center Kyoto Prefectural University of Medicine, Kyoto Yamashiro General Medical Center, Kyorin University Hospital, Kiryu Kosei General Hospital, Kindai University Nara Hospital, Kushiro Rosai Hospital, Japanese Red Cross Kumamoto Hospital, Kurume University Hospital, Kurobe City Hospital, Gunma Children’s Medical Center, Gunma Prefectural Cancer Center, Keio University Hospital, Kesen-numa City Hospital, Kanoya Medical Center, Kochi Medical School Hospital, Kinki Central Hospital of the Mutual Aid Association of Public School Teachers, Koga General Hospital, Tosei General Hospital, Naga Hospital, Yame General Hospital, Fujioka General Hospital, National Cerebral and Cardiovascular Center Hospital, National Hospital Organization Kanazawa Medical Center, National Hospital Organization Kofu National Hospital, National Hospital Organization Kobe Medical Center, National Hospital Organization Chiba Medical Center, National Hospital Organization Saitama Hospital, National Hospital Organization Saga Hospital, Komaki City Hospital, Saiseikai Sendai Hospital, Saiseikai Niigata Hospital, Yamagata Saisei Hospital, Saitama Medical University Hospital, Saitama Medical Center, Saitama City Hospital, Saga Prefectural Hospital Koseikan, Shiga General Hospital, National Hospital Organization Shikoku Cancer Center, Shizuoka Children’s Hospital, Shizuoka Cancer Center, Shimane Prefectural Central Hospital, Shimane University Hospital, San-Ikukai Hospital, Tagawa Hospital, Juntendo University Shizuoka Hospital, Shobara Redcross Hospital, Shirakawa Kosei General Hospital, Uwajima City Hospital, Kishiwada City Hospital, Kofu City Hospital, Municipal Tsuruga Hospital, Toyonaka Municipal Hospital, Nagahama City Hospital, Wakkanai City Hospital, Tonami General Hospital, Shin-Oyama City Hospital, National Hospital Organization Shinshu Ueda Medical Center, Suzuka General Hospital, Suwa Red Cross Hospital, Sendai Red Cross Hospital, Sendai City Hospital, Aiiku Hospital, Takaoka City Hospital, National Hospital Organization Takasaki General Medical Center, Takatsuki General Hospital, Tanabe Chuo Hospital, Chugoku Rosai Hospital, Tsuchiura Kyodo General Hospital, Tsuyama Chuo Hospital, Tsuruoka Municipal Shonai Hospital, National Hospital Organization Tsuruga Medical Center, Tenri Hospital, Tokyo Medical University Ibaraki Medical Center, Tokyo Medical University Hospital, The University of Tokyo Hospital, Tokyo Metropolitan Ohtsuka Hospital, Tokyo Metropolitan Komagome Hospital, KKR Tohoku Kosai Hospital, Tokushima Prefectural Central Hospital, Tokushima Red Cross Hospital, National Hospital Organization Okayama Medical Center, National Hospital Organization Hirosaki National Hospital, National Hospital Organization Fukuoka-higashi Medical Center, Tochigi Cancer Center, Dokkyo Medical University Saitama Medical Center, Tottori Pref. Kousei Hospital, Tottori University Hospital, Rumoi City Hospital, Toyama City Hospital, Toyama University Hospital, Toyama Rosai Hospital, TOYOTA Memorial Hospital, Toyota Kosei Hospital, Toyohashi Municipal Hospital, Nagaoka Chuo General Hospital, National Hospital Organization Nagasaki Medical Center, Nagano Children’s Hospital, Nagara Medical Center, Nagoya Daini Hospital, National Hospital Organization Nagoya Medical Center, Nasu Red Cross Hospital, Nayoro City General Hospital, Nara Medical University Hospital, Nara Prefecture General Medical Center, Japanese Red Cross Narita Hospital, Niigata Cancer Center Hospital, Niigata Prefectural Shibata Hospital, Niigata Prefectural Central Hospital, Niigata City General Hospital, Nishiwaki Municipal Hospital, Nippon Medical School Musashikosugi Hospital, Nihonkai General Hospital, Nihon University Itabashi Hospital, The Japan Baptist Hospital, Numazu City Hospital, Noshiro Kousei Medical Center, Hamamatsu Medical Center, Hannan Chuo Hospital, National Hospital Organization Higashihiroshima Medical Center, Nikko Memorial Hospital, Hitachi General Hospital, Hitoyoshi Medical Center, Japanese Red Cross Society Himeji Hospital, Hyogo Prefectural Awaji Medical Center, Hyogo Prefectural Kobe Children’s Hospital, Hyogo Prefectural Kaibara Hospital, Hiraka General Hospital, Hiroshima University Hospital, Fukagawa Municipal Hospital, Fukaya Red Cross Hospital, Fukui Aiiku Hospital, Fukui Prefectural Hospital, Fukui General Hospital, Fukuoka University Hospital, Fukuyama City Hospital, Fujieda Municipal General Hospital, Fuji City General Hospital, Fujimoto General Hospital, Funabashi Central Hospital, Funabashi Municipal Medical Center, Furano Kyokai Hospital, Beppu Medical Center, Maizuru Kyosai Hospital, Masuda Red Cross Hospital, Matsudo City General Hospital, Matsuyama Red Cross Hospital, Matsusaka Chuo General Hospital, Mie Chuo Medical Center, Mie Prefectural General Medical Center, Misawa Municipal Misawa Hospital, Japanese Red Cross Mito Hospital, National Hospital Organization Minami Wakayama Medical Center, South Miyagi Medical Center, Miyagi Cancer Center, Miyagi Children’s Hospital, Mutsu General Hospital, Yaizu City Hospital, Yao Municipal Hospital, Yakumo General Hospital, Yamagata University Hospital, Yamagata Prefectural Shinjo Hospital, Yamagata Prefectural Central Hospital, Syuutou General Hospital, Yamaguchi-ken Saiseikai Shimonoseki General Hospital, Yokosuka Kyosai Hospital, Yokohama Municipal Citizen’s Hospital, Haga Red Cross Hospital, Rinku General Medical Center, Wakayama Medical University Hospital, Imakiire General Hospital, Teine Keijinkai Hospital.

