## Supporting Information for "Trends in pregnancy-associated cervical cancer in Japan between 2012 and 2017: A multicenter survey"

**Figure S1. Annual trends in the frequencies of cervical cancer during pregnancy and postpartum.**

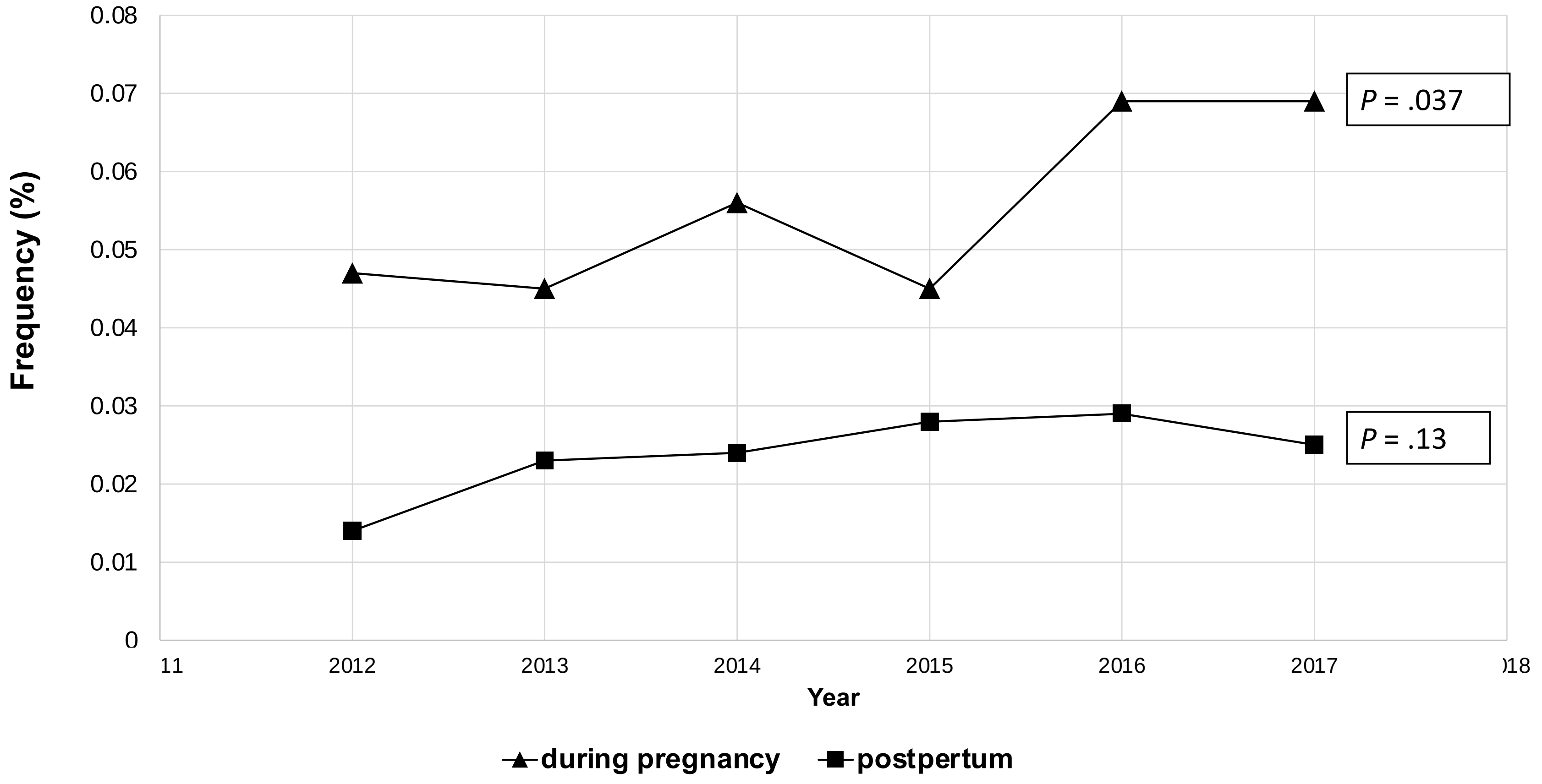

**Figure S2. Time from diagnosis to delivery in the group diagnosed at ≥22 weeks of gestation**

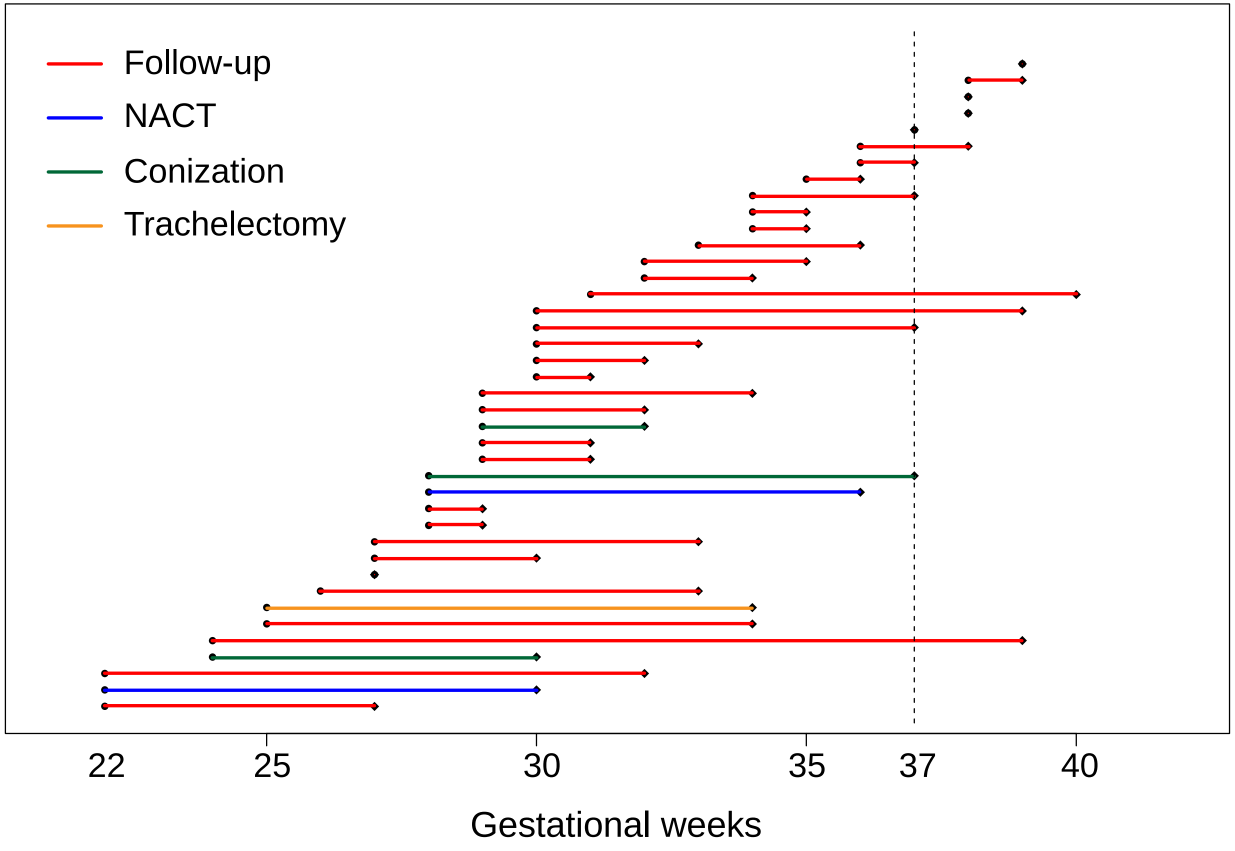

**Figure S3. Pregnancy outcome of stage IA1 cases treated by conization before <22 weeks of gestation.**

**Duration from diagnosis to delivery (A) and the median gestational age at diagnosis and delivery (B) in stage IA1 cases who underwent conization before <22 weeks of gestation were**

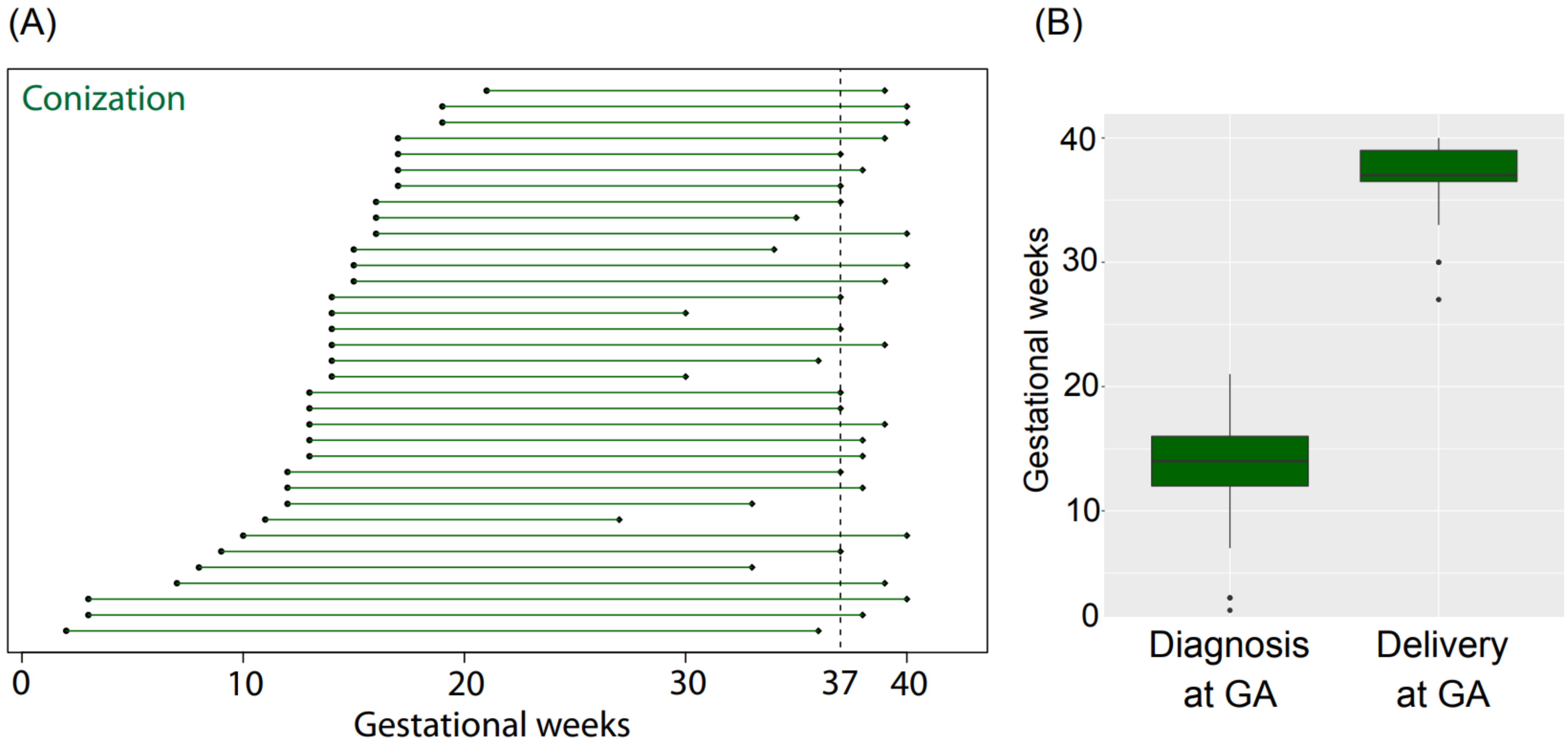

**Figure S4. Subsequent treatments in the four groups**

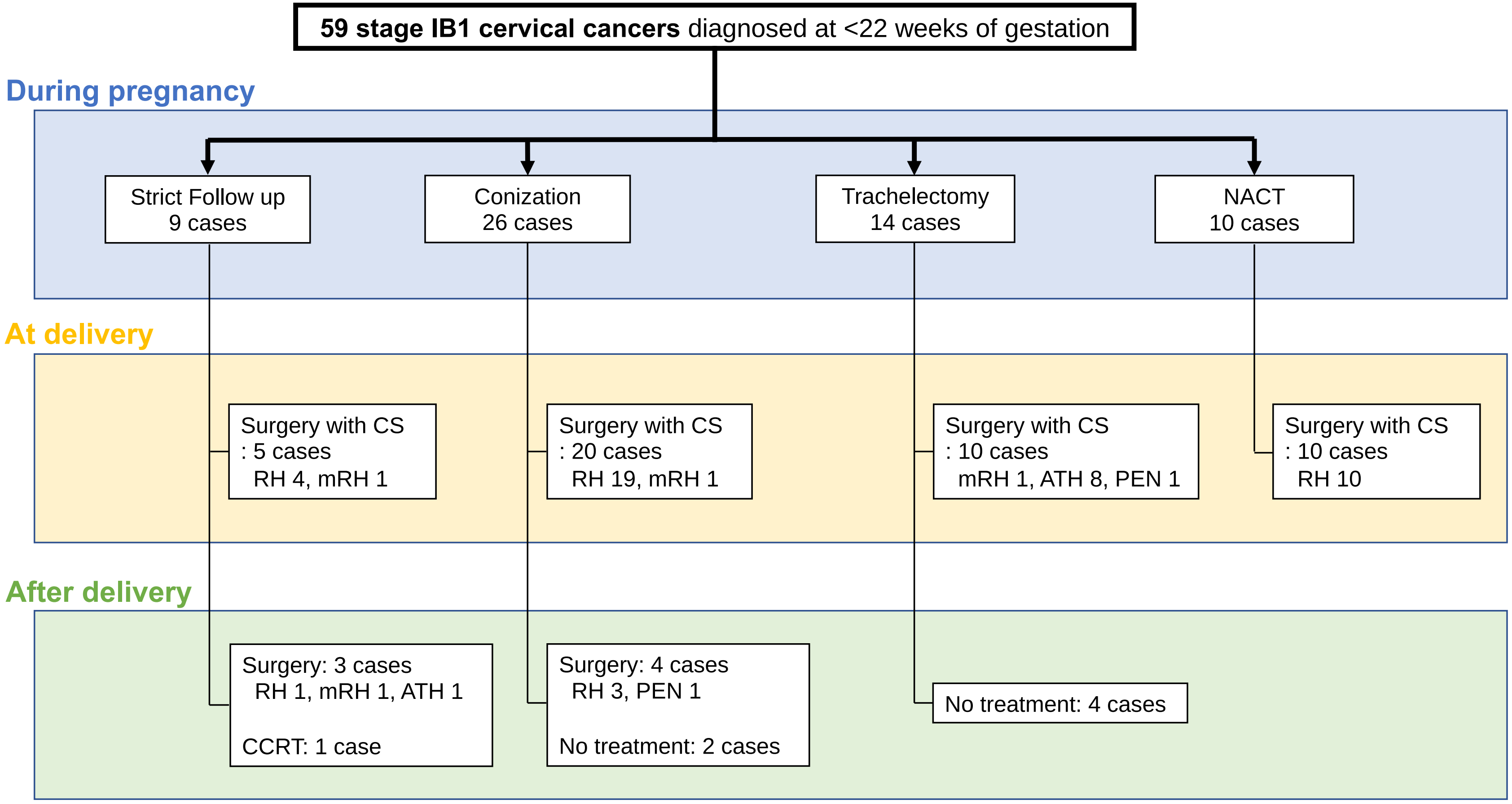


ATH: Simple hysterectomy, CCRT: concurrent chemoradiotherapy, CS: cesarian section, mRH: modified radical hysterectomy,

NACT: neoadjuvant chemotherapy, PEN: pelvic lymphadenectomy, RH: Radical hysterectomy

**Table S1. Trends in treatment modalities administered during pregnancy by stage among patients diagnosed at a gestational age of ≥22 weeks (n=40)**

| **Treatment modality** | **IA1 (n=3)** | **IA2** (n = 1) | **IB1** (n = 20) | **IB2- IVB** (n = 16) |
|---|---|---|---|---|
| Strict follow up (n = 34) | 2 (66.7%) | 1 (100%) | 17 (85.0%) | 14 (87.5%) * |
| Conization (n = 3) | 1 (33.3%) | - | 2 (10.0%) | - |
| Trachelectomy (n = 1) | - | - | 1 (5.0%) | - |
| NACT (n = 2) | - | - | - | 2 (12.5%) |

Data are presented as n (%).

* Three patients died of disease
